# Increased Self-Reported Discrimination and Concern for Physical Assault Due to the COVID-19 Pandemic in Chinese, Vietnamese, Korean, Japanese, and Filipino Americans

**DOI:** 10.1101/2020.09.15.20194720

**Authors:** Sierra K. Ha, Ann T. Nguyen, Chloe Sales, Rachel S. Chang, Hillary Ta, Malathi Srinivasan, Sukyung Chung, Latha Palaniappan, Bryant Lin

## Abstract

**Objectives:** To investigate self-reported discrimination and concern for physical assault due to the COVID-19 pandemic among disaggregated Asian subgroups in the US.

**Methods:** We conducted a nationwide survey to assess self-reported discrimination and concern for physical assault due to COVID-19 across racial/ethnic groups, including diverse subgroups of Asians.

**Results:** Chinese respondents experienced the largest change (15% increase) in proportion of respondents reporting discrimination from 2019 to 2020 (P<.01). Chinese, Korean, Japanese, Vietnamese, and Other API showed up to 3.9 times increased odds of self-reported racial/ethnic discrimination due to COVID-19 and, with the addition of Filipino, experienced up to 5.4 times increased odds of concern for physical assault due to COVID-19 compared to Whites.

**Conclusions:** Our study is the first to examine self-reported discrimination and concern for physical assault due to COVID-19 in subgroups of Asian Americans, finding that East (Chinese, Korean, Japanese) and Southeast (Vietnamese, Filipino) Asian Americans have been disproportionately affected. Future studies should disaggregate Asian subgroups to fully understand experiences of discrimination in diverse populations in the US.

## INTRODUCTION

The COVID-19 pandemic has polarized America and reports of discrimination have increased in many racial groups.^1–3^ Notably, public figures and the media have promoted racist rhetoric implicating Chinese in the pandemic, coining terms such as the “Chinese virus” or “Kung Flu.”^4,5^ Several sources have reported that the popularization of these views has resulted in increased racist and xenophobic discrimination.^6–8^ Many incidents have targeted not only people of Chinese descent but also the broader population of people of any Asian descent.^9^ During the pandemic from March 19 to August 5, 2020, one advocacy center, STOP AAPI HATE, received reports of over 2,583 incidents of discrimination ranging from verbal harassment to physical assault against Asian Americans across the United States.^6^ Other experiences of Asians during the pandemic have included vandalization, being refused service, and decreased patronization of Asian-owned businesses at the start of the pandemic.^6,9,10^

Despite the increase in anti-Asian discriminatory incidents, few studies have examined the prevalence and effects of these experiences on Asian Americans. A recent study found that since the outbreak, Asians are twice as likely to experience discrimination compared to Whites, which was associated with significant mental distress.^11^ Asians are also shown to be more likely to report adverse experiences due to their race or ethnicity, with three in ten Asian adults being subjected to racial slurs or jokes.^2^ However, these studies were conducted on small Asian American sample sizes (278 and 119 respectively) and aggregated Asian American and Pacific Islander ethnic subgroups. Given the heterogeneity of Asians in the United States, disaggregating this data is vital to unmask the effects of discrimination on Asian American subgroups and address racial injustice in diverse populations as a matter of public health.^12^

We conducted a nationwide survey to understand the prevalence and severity of self-reported racial/ethnic discrimination due to the COVID-19 pandemic in the United States across disaggregated Asian subgroups, Whites, Hispanics/Latinos, and Blacks. We specifically assessed self-reported race/ethnicity, participant place of birth, mental distress, self-reported discrimination and concern for physical assault due to COVID-19. To our knowledge, this study is the largest survey to assess the impact of COVID-19 discrimination on disaggregated Asian subgroups in the United States.

## METHODS

### IRB

The study was considered exempt by Stanford University Institutional Review Board (Protocol: 56235)

### Funding

The study was funded by the Stanford Center for Asian Health Research and Education.

### Study design

We conducted a nationwide online survey on self-reported discrimination using convenience sampling. The survey was distributed via Pollfish and email listservs for Asian American organizations in order to achieve an oversampling of Asian Americans.

### Main Measures

Survey Development: Based on prior survey instruments^11,13,14^, we developed a survey matrix to explore the major domains of discrimination, including being hassled or made to feel inferior, physical violence, and mental distress specifically in relation to the COVID-19 pandemic. The survey questions were designed to include wording that had clear translations in Asian languages. Translation was performed by Interpretation and Translation Services at Stanford Health, and back translation was performed by an independent translator to ensure that exact meaning was conveyed. An initial 20 item survey was pilot tested with Stanford Medicine faculty and student interest groups. We revised the survey after feedback to 15 final items (Supplemental Materials 1), including demographics (age, gender, educational level, immigration status), ethnicity (White; Hispanic or Latino; Black; Asian American or Pacific Islander: Chinese, Vietnamese, Korean, Japanese, Filipino, South Asian, Other API; Other), change in behavior due to the COVID-19 pandemic, self-reported discrimination overall (2019 and 2020), self-reported racial/ethnic discrimination due to COVID-19, physical assault due to COVID-19 (concern and actual), and mental health (Patient Health Questionnaire-4).

We assessed self-reported discrimination as being “hassled or made to feel inferior because of race or ethnicity.” To measure self-reported discrimination overall, Participants were asked “Were you ever hassled or made to feel inferior because of your race or ethnicity for any reason?” for both 2019 and 2020. Participants answered “yes”, “no”, or “unsure.” To measure self-reported racial/ethnic discrimination due to COVID-19, participants were asked “Were you ever hassled or made to feel inferior because of your race or ethnicity as related to the COVID-19 disease?” Participants responded “yes”, “no”, or “unsure”.

To measure COVID-19 related physical assault, participants were asked, “In the year 2020, were you ever physically assaulted due to your race or ethnicity as related to the COVID-19 disease?”. To differentiate between actual incidents of assault and concern for COVID-19 related physical assault, participants were asked, “In the year 2020, were you ever concerned about physical assault due to your race or ethnicity as related to the COVID-19 disease?” Both questions required participants to answer “yes”, “no”, or “unsure.”

The survey was implemented through Qualtrics (Provo, Utah) and Pollfish (New York City, New York).

### Participants

The survey was conducted in two phases, with the survey in the second phase including an additional question on immigration status. In both phases, we recruited participants via two simultaneous approaches: (1) Pollfish, a paid online survey platform with a pre-recruited sample^15^ and (2) a Qualtrics survey link distributed via social media and email listservs identified as serving Asian American groups. For the latter, an advertising flyer explained the study and encouraged participants to respond to the survey with the provided link. The flyer and survey were made available in multiple languages, including English, Chinese (Traditional), Chinese (Simplified), Korean, Vietnamese, and Tagalog. Outreach for the first phase began on May 13th, 2020 and concluded on July 19th, 2020. The second phase of the survey, including the question on immigration status, was distributed via the same approaches from July 20th, 2020 to August 12, 2020. After excluding 95 participants who did not answer all survey questions, a total of 1,861 responses were obtained over both phases, with 1,588 from Pollfish and 273 from

Qualtrics. Pollfish participants received a small non-cash incentive through the platform in exchange for a completed survey^15^, whereas no incentive was provided for all other respondents who completed the Qualtrics survey. Respondents came from 50 states and the District of Columbia, with similar proportions of total state population from 2010 US Census.^16^

### Analysis

Of 1861 respondents that completed the survey, 1,038 were from Phase 1 (1004 Pollfish and 34 Qualtrics) and 823 were from Phase 2 (584 Pollfish and 239 Qualtrics). After 44 participants who listed their race/ethnicity as “Other” were excluded, a total of 1817 questionnaires were analyzed. We categorized responses to discrimination and physical assault measures as “yes” or “no” and “unsure,” and participants who answered “no” or “unsure” served as the reference group for those who answered “yes.” Less than 9% of participants responded “unsure” to each variable. We examined the difference in self-reported discrimination overall before (2019) and during (2020) the pandemic across all ethnic groups, using chi-squared testing for overall difference and pairwise comparisons across racial/ethnic groups. We then used multivariate logistic regressions of self-reported racial/ethnic discrimination due to COVID-19 with the key predictor of race/ethnicity, using White as the reference group, and covariates including age, gender, education level, geographic location, self-reported discrimination overall in 2019, and survey phase. Similarly, a multivariable logistic model was used for physical assault on the same set of covariates. To take into account multiple comparisons, we considered P<.01 as statistically significant, and reported 99% confidence intervals. Analyses were performed using RStudio (Version 1.2.5001, Boston, MA).

## RESULTS

Table 1 shows the demographic profile of our respondents based on race/ethnicity, age, gender, education attainment, and immigration status. The Asian American and Pacific Islanders were further stratified to Chinese (25%), Korean (6%), Japanese (6%), Vietnamese (9%), Filipino (14%), and South Asian (which includes Indian and Pakistani – 18%). Race/ethnicity was not independent from age, gender, immigration, and education by chi-squared analysis, so these demographic variables were adjusted for in subsequent multivariate models.

**TABLE 1.** — Participant Characteristics by Self-Reported Race/Ethnicity: United States, 2020. *Note*. Testing of difference was conducted using chi-square test. Percentages may not add up to 100% as percentages were rounded. *Immigration status was asked in the second phase of the survey only.

|  | White<br>(n = 295)<br>No. (%) | Chinese<br>(n = 333)<br>No. (%) | Korean<br>(n = 83)<br>No. (%) | Japanese<br>(n = 77)<br>No. (%) | Vietnamese<br>(n = 124)<br>No. (%) | Filipino<br>(n = 187)<br>No. (%) | South Asian<br>(n = 241)<br>No. (%) | Other API<br>(n = 280)<br>No. (%) | Hispanic or<br>Latino<br>(n = 142)<br>No. (%) | Black<br>(n = 55)<br>No. (%) | Total<br>(n = 1817)<br>No. (%) | P |
| --- | --- | --- | --- | --- | --- | --- | --- | --- | --- | --- | --- | --- |
| Age |  |  |  |  |  |  |  |  |  |  |  | <.001 |
| 18 - 24 | 48 (16) | 98 (29) | 19 (23) | 6 (7) | 54 (43) | 16 (9) | 42 (17) | 107 (38) | 34 (24) | 8 (145) | 432 (24) |  |
| 25 - 34 | 86 (29) | 76 (23) | 23 (28) | 13 (17) | 24 (19) | 39 (21) | 54 (22) | 72 (26) | 40 (28) | 12 (22) | 439 (24) |  |
| 35 - 44 | 69 (23) | 81 (24) | 26 (31) | 18 (23) | 23 (19) | 75 (40) | 90 (37) | 71 (25) | 43 (30) | 21 (38) | 517 (29) |  |
| 45 - 54 | 44 (15) | 41 (12) | 9 (11) | 19 (25) | 12 (10) | 43 (23) | 33 (14) | 17 (6) | 20 (14) | 8 (15) | 246 (14) |  |
| Over 54 | 48 (16) | 37 (11) | 6 (7) | 21 (27) | 11 (9) | 14 (8) | 22 (9) | 13 (5) | 5 (4) | 6 (11) | 183 (10) |  |
| Female | 160 (54) | 193 (58) | 43 (52) | 36 (47) | 65 (52) | 132 (71) | 135 (56) | 155 (55) | 82 (58) | 20 (36) | 1021 (56) | <.001 |
| Education |  |  |  |  |  |  |  |  |  |  |  | <.001 |
| No College | 79 (27) | 48 (14) | 7 (8) | 9 (12) | 26 (21) | 32 (17) | 33 (14) | 72 (26) | 47 (33) | 18 (33) | 371 (20) |  |
| College or Higher | 216 (73) | 285 (86) | 76 (92) | 68 (88) | 98 (79) | 155 (83) | 208 (86) | 208 (74) | 95 (67) | 37 (67) | 1446 (80) |  |
| Immigration* (n = 817) |  |  |  |  |  |  |  |  |  |  |  | <.001 |
| Born in the US | 67 (97) | 104 (54) | 23 (55) | 29 (64) | 28 (64) | 34 (29) | 35 (27) | 77 (69) | 30 (94) | 30 (91) | 457 (56) |  |
| Moved to the US before age 18 | 0 (0) | 45 (24) | 15 (36) | 8 (18) | 12 (27) | 24 (20) | 18 (14) | 13 (12) | 0 (0) | 3 (9) | 138 (17) |  |
| Moved to the US age 18 or later | 2 (3) | 42 (22) | 4 (9) | 8 (18) | 4 (9) | 61 (51) | 78 (60) | 21 (19) | 2 (6) | 0 (0) | 222 (27) |  |

Among all respondents, Chinese respondents experienced the highest increase in self-reported discrimination overall from 2019 to 2020 (15%, P<.01). The increases in Vietnamese (13%) and Other API (9%) were larger than in White (7%), but these differences were not statistically significant (Figure 1).

**FIGURE 1.**
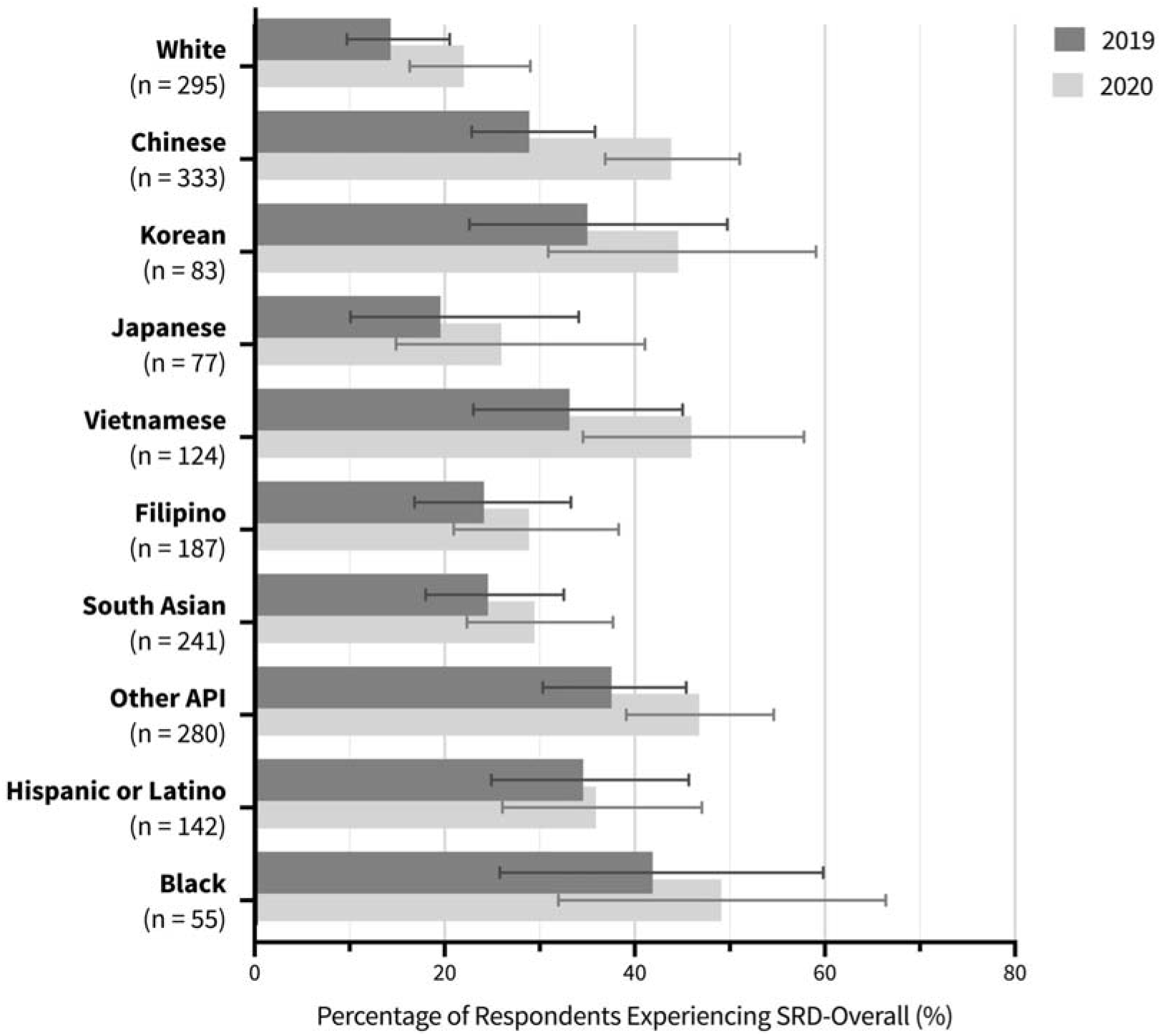
—Percentage of Respondents Experiencing Self-Reported Discrimination Overall by Race/Ethnicity: United States, 2019-2020. ***Note***. Chi-squared analysis shows a significant change in the proportion of Chinese respondents who reported self-reported discrimination from 2019 to 2020 (P<.01). All other race/ethnic groups did not experience a significant change in self-reported discrimination 2019 to 2020.

Vietnamese (58%), Chinese (51%) and Other API (43%) respondents had the highest proportion of reported concern for physical assault due to COVID-19 compared to White respondents (all P<.001). Filipino (32%) and Korean (41%) respondents also had a higher proportion of reported concern for physical assault compared to White respondents (all P<.01). South Asian, Black and Hispanic or Latino respondents did not differ from White in the proportion of concern for physical assault (Supplemental Figure 1).

Of Asian American subgroups, Vietnamese were most likely (46%) to self-report racial/ethnic discrimination due to COVID-19, followed by Koreans (43%), and South Asians were least likely to report incidents of perceived discrimination (12%). The proportion of self-reported racial/ethnic discrimination due to COVID-19 was the highest for Vietnamese, followed by Korean, Chinese, and Other API, all of which were greater than White (all P<.01). The proportion of reported incidents of perceived discrimination did not significantly differ in Japanese, Filipino, South Asian, Black, and Hispanic respondents compared to White respondents (Supplemental Figure 1).

After adjusting for age, gender, education, prior discrimination experience, and the survey distribution phase, Chinese (AOR = 3.5, 99% CI = 1.9, 6.5), Korean (AOR = 3.8, 99% CI = 1.7, 8.4), Japanese (AOR = 3.1, 99% CI = 1.3, 7.5), Vietnamese (AOR = 3.9, 99% CI = 1.9, 8.0), and Other API (AOR = 2.7, 99% CI = 1.5, 5.0) respondents were more likely to self-report racial/ethnic discrimination due to COVID-19 compared to White respondents, but South Asian and Filipino respondents did not differ from White respondents in likelihood of experiencing discrimination (Figure 2).

**FIGURE 2.**
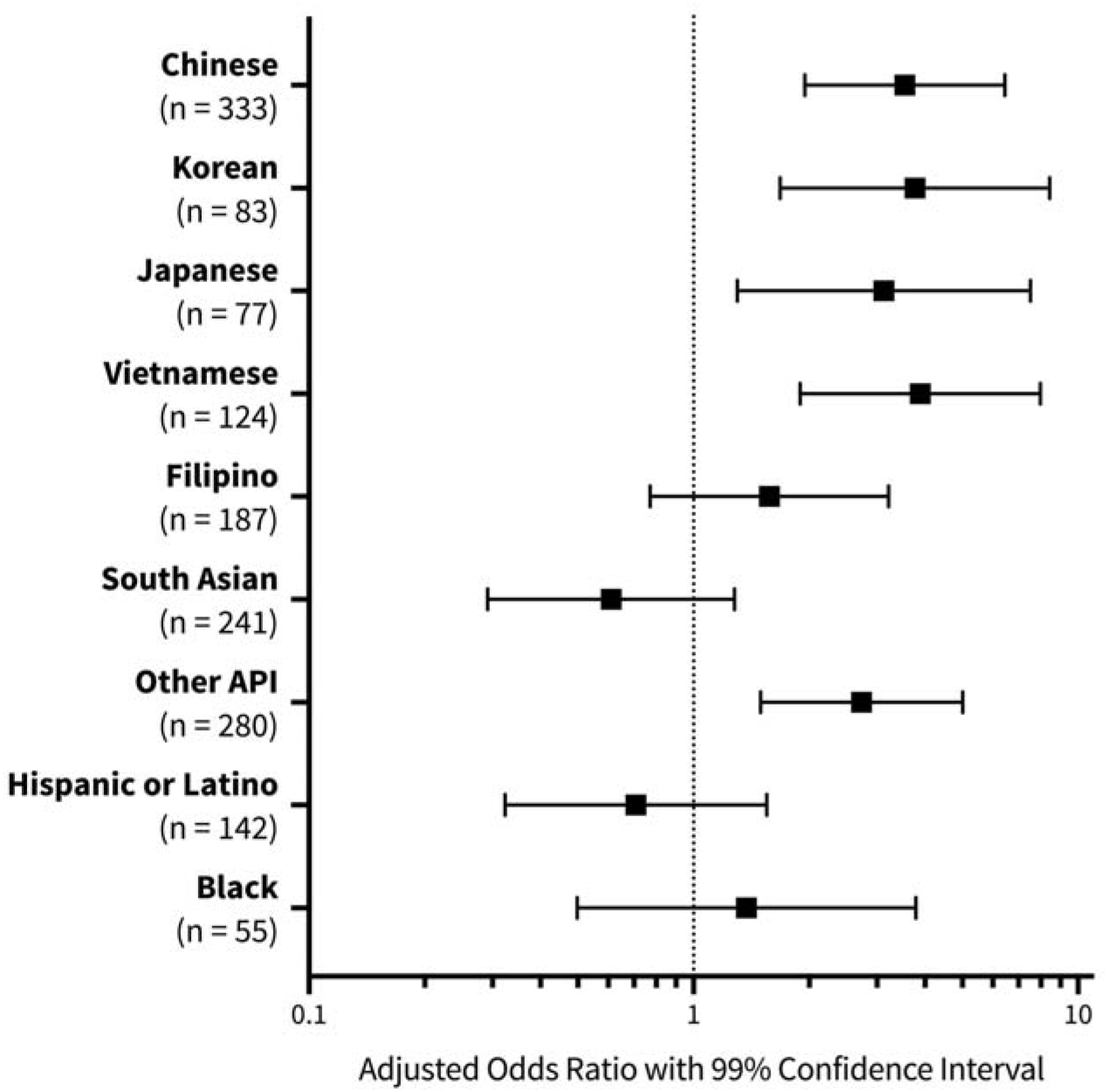
—Adjusted Odds Ratios of Self-Reported Racial/Ethnic Discrimination due to COVID-19 by Race/Ethnicity: United States, 2020. *Note*. Odd ratios adjusted for age, gender, education, prior discrimination, and survey distribution phase (1 or 2), compared to Whites.

The results for concern for physical assault due to race/ethnicity related to the COVID-19 pandemic were similar to those of self-reported racial/ethnic discrimination due to COVID-19, such that Chinese (AOR = 4.4, 99% CI = 2.6, 7.7), Vietnamese (AOR = 5.4, 99% CI = 2.8, 10.6), Japanese (AOR = 2.3, 99% CI = 1.0, 5.3) and Other API (AOR = 2.4, 99% CI = 1.4, 4.3) respondents were more likely to experience this concern than White respondents. Additionally, Filipino (AOR = 2.2, 99% CI = 1.2, 4.1) respondents were also more likely to report concern for physical assault due to race/ethnicity related to COVID-19 than White respondents (Figure 3). In addition to race/ethnicity, age and prior experience of discrimination were associated with increased odds of both self-reported racial/ethnic discrimination due to COVID-19 and concern for physical assault due to COVID-19 (Supplemental Table 1). Mental distress, as measured by PHQ-4, was not higher in Asian Americans compared to Whites, despite higher self-reported racial/ethnic discrimination in Asian Americans.

**FIGURE 3.**
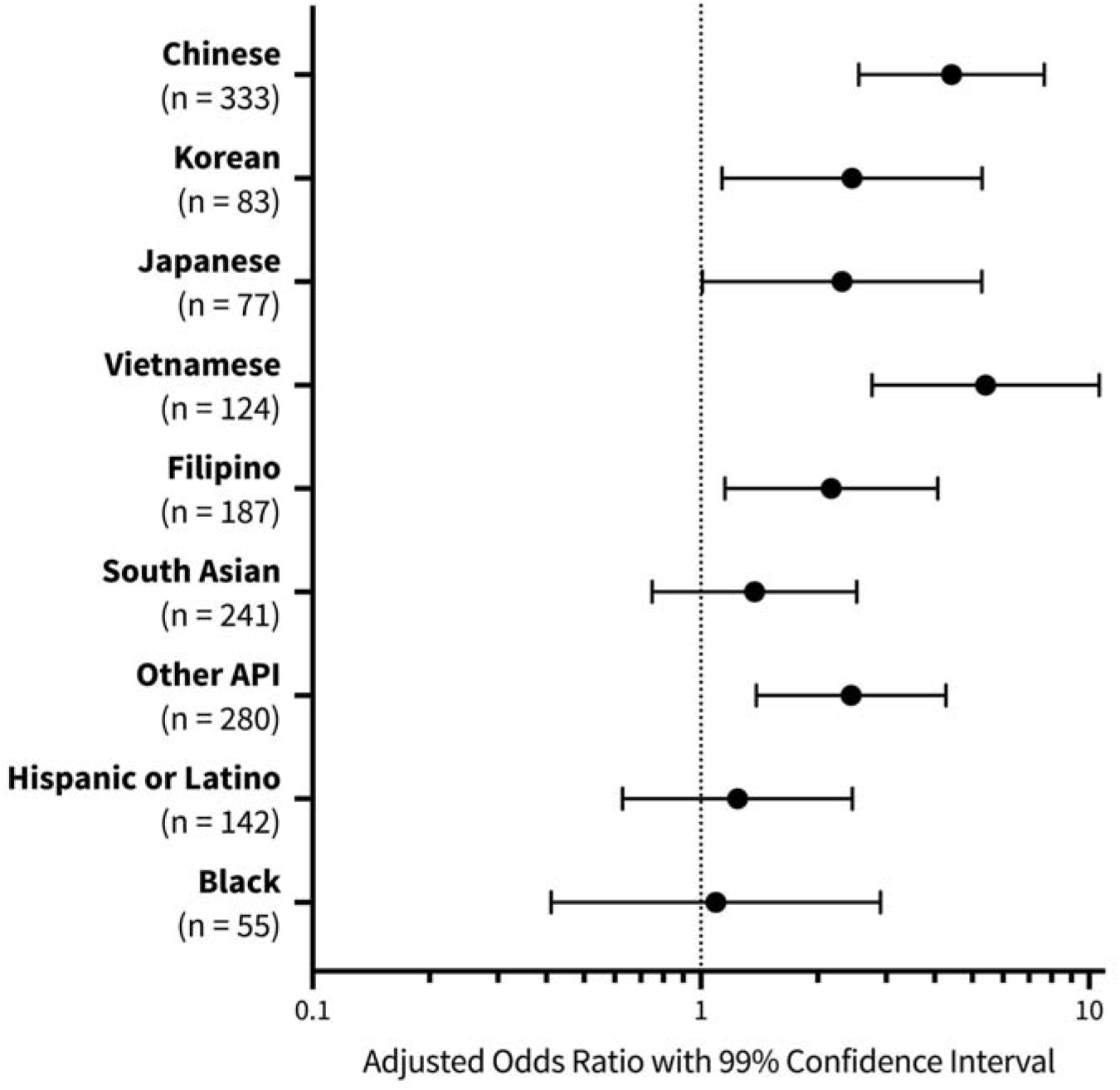
—Adjusted Odds Ratios of Concern for Physical Assault due to COVID-19 by Race/Ethnicity: United States, 2020. *Note*. Odd ratios adjusted for age, gender, education, prior discrimination, and survey distribution phase (1 or 2), compared to Whites.

## DISCUSSION

The current study sought to examine self-reported discrimination in Asian American subgroups during the COVID-19 pandemic. Our findings show that Chinese Americans have experienced a significant increase in overall racial/ethnic self-reported discrimination from before (2019) to during (2020) the pandemic. Chinese, Korean, Japanese, Vietnamese, Filipino, and Other API subgroups have been adversely affected by pandemic, as measured by concern for physical assault and self-reported racial/ethnic discrimination due to the COVID-19 pandemic, in comparison to Whites. Our findings are the first to oversample Asian Americans, disaggregate Asian subgroups, and provide a national cross section of perceived discrimination, giving insight into the domains of discrimination faced especially by East (Chinese, Korean, Japanese) and Southeast (Vietnamese, Filipino) Asians during the pandemic.

Our findings reveal that Asian Americans have been adversely affected by the COVID-19 pandemic, supporting the findings of previous studies and anecdotal evidence of anti-Asian violence, assault and discriminatory attitudes towards these groups.^2,3,6,9,11^ However, certain Asian American subgroups experienced higher self-reported racial/ethnic discrimination due to COVID-19 (up to 3.9 times higher in Vietnamese Americans) compared with Whites, suggesting that the issue may be more severe than previous findings that Asian Americans as a group were 2 times more likely to experience COVID-19 associated discrimination during April 2020.^11^ Chinese, Korean, Japanese, Vietnamese, and Other API groups in the current study all experienced increased self-reported racial/ethnic discrimination due to COVID-19 than the rates previously reported for Asian Americans overall.^11^ Additionally, while the current study found that certain Asian American subgroups reported high concerns for physical assault (up to 46% in Vietnamese Americans), previous studies have found that only 26% of Asian Americans overall feared someone might threaten or physically attack them.^2^ These prior studies^2,11^ did not disaggregate by Asian ethnic subgroups, masking any possible differences in experiences between them. Additionally, their small sample size of 119 and 278 Asian respondents, respectively, compared to the current sample of 1325 Asian American respondents could have contributed to these observed differences in effect size between studies.

The current study found that during the pandemic, East Asians, including Chinese, Korean, and Japanese, have frequently been targeted in discriminatory incidents and have expressed high levels of concern for physical assault, which can be rationalized due to the fact that these groups share more facial and physical similarities with Chinese^17^, who have been linked to the coronavirus outbreak. Southeast Asians in our study (Vietnamese, Filipino) were also more likely to experience discrimination and express concern for physical assault, while South Asians did not differ significantly in their responses compared to Whites. The disparities in prevalence and magnitude of these experiences among different Asian American ethnic subgroups, would be expected given the heterogeneous socioeconomic and cultural characteristics and history of discrimination among Asian Americans.^18–20^ The differences among Asian American subgroups in this study illustrates the need for better approaches to studying Asian American populations throughout research, such as disaggregation of data on Asian Americans, especially in studies focusing on the diverse Asian American population in the US.^12,21,22^ Future studies should further explore this approach in order to assess the impact of the pandemic on the Asian American population.

Chinese Americans have faced increased anti-Asian xenophobia and racism because of the association of China with COVID-19.^8,23^ The proportion of Chinese Americans experiencing any type of discrimination has increased by over 50% over the course of the pandemic. While other Asian American groups (Korean, Japanese, Vietnamese, Filipino, South Asian, Other API) did report changes in discrimination from before (2019) to during (2020) the pandemic, these increases were not statistically significant, which could be attributed to the higher baseline discrimination (2019) faced by some of these groups (Korean, Vietnamese, Other API) compared to Chinese Americans.

After adjusting for age, gender, education, prior discrimination experience, and the survey distribution phase, Vietnamese Americans were more than 3 times more likely to experience self-reported racial/ethnic discrimination due to COVID-19 and 5 times more likely to be concerned about physical assault than Whites, the highest likelihood seen amongst all Asian American subgroups in the current study. This contrasts previous findings that Vietnamese are less likely to perceive discrimination than Chinese and Korean Americans.^18^ Given historical conflicts and the recent increase in tension between the nations of Vietnam and China over the South China Sea^24,25^, one possible reason for this observation is that Vietnamese respondents may have been especially angered by being associated with China, which has been blamed for the coronavirus, and having to bear the same burden of discrimination as Chinese Americans. Future qualitative work in Vietnamese Americans may be necessary to fully contextualize these early findings.

While the current study presented robust evidence for the experiences of Asian Americans related to COVID-19, several limitations should be considered in the interpretation of these findings. Firstly, our use of a novel measurement of self-reported racial/ethnic discrimination due to COVID-19 has not been validated. The survey, which was advertised and administered via online platforms including email listservs, social media, and Pollfish, limited our sample to technologically literate populations, leading to a lack of older respondents (Table 1) and relatively few respondents with limited English proficiency, despite the fact that alternative language surveys were offered. Additionally, given the cross-sectional, retrospective, and survey nature of our current study, selection, recall, and social desirability biases may have all played a role in overrepresenting or underrepresenting participants experiencing discrimination. There is no evidence to suggest that these biases are differential by race/ethnicity, though these racial/ethnic comparisons should be viewed with these caveats in mind. While this is the largest sample of COVID-19 discrimination survey data in Asian Americans published to date, each Asian subgroup displayed a relatively small sample size. Future studies should strive to disaggregate Asian subgroups and achieve higher sample sizes to further explore trends that were observed in our study. There are also numerous factors not controlled for in this study that could have contributed to participant self-reported discrimination, including exposure to social media and differences in shelter-in-place orders across regions.^11,26^

While the current study did not show an increase in mental distress associated with self-reported racial/ethnic discrimination due to COVID-19 among Asian subgroups, previous studies have shown that Asians are less likely to report anxiety and depression because of stigma attached to mental disorders.^27–29^ Despite the fact that an increase in anxiety and depression was not shown in this study, future studies should further explore the possible implications of COVID-19 discrimination on health and wellness using instruments that are sensitive in Asian populations.^30^

The findings of this study illustrate the differences in discrimination experienced across diverse Asian American communities. This study confirms anecdotal evidence and builds on prior studies on Asian Americans as a group that have explored discrimination.^2,9,11^ Our study suggests that the anti-Chinese sentiment^5,23,31^ related to the COVID-19 pandemic has affected not only Chinese Americans, but also a wide range of other Asian Americans including East Asians (Japanese, Korean) and Southeast Asians (Vietnamese, Filipino). Including less affected groups (South Asians) in aggregate with these more affected groups may underestimate overall estimates of COVID-19 related discrimination among Asian Americans as a group. Compounded with the social and health challenges that the pandemic has brought upon all Americans,^26^ this surge in harmful xenophobia and racism towards Asian American populations related to the COVID-19 pandemic represents a major public health issue in the United States requiring further study and intervention efforts.

## CONTRIBUTORS

B. Lin, L. Palaniappan, and C. Sales conceptualized the study. S. Ha and A. Nguyen led the data collection, analysis, and writing. All of the authors contributed to interpreting the results and drafting the article.

## Data Availability

The datasets generated during and/or analyzed during the current study are not publicly available due to their containing information that could compromise the privacy of research participants but are available from the corresponding author on reasonable request.

## ACKNOWLEDGEMENTS

This project was supported by the Stanford Center for Asian Health Research and Education (CARE). We thank the National Board of the Asian Pacific American Medical Student Association (APAMSA, http://www.apamsa.org) for their invaluable assistance with disseminating our Qualtrics survey through their national channels. We also thank the leadership of the Stanford Medicine Chapter of APAMSA: Vivian Lou, Richard Liang, and Huiying “Becky” Tsang.

## HUMAN PARTICIPANT PROTECTION

This study was approved by Stanford University’s Research Compliance Office. Participants were provided information about the study and were considered to have provided informed consent by continuing to complete the anonymous survey.

